# China’s fight against COVID-19: What we have done and what we should do next?

**DOI:** 10.1101/2020.03.28.20046086

**Authors:** Sixiang Cheng, Yuxin Zhao, Atipatsa Chiwanda Kaminga, Pingxin Zhang, Huilan Xu

## Abstract

**Background:** On 12 March, the World Health Organization Director-General declared that “the threat of a global pandemic has become a reality”, and the disease caused by the novel coronavirus, known as COVID-19, has become a global concern. Chinese efforts in curbing the virus have widely been recognized. Even the WHO has lauded the efforts of the Chinese government and advised the world to learn from China in fighting the disease. Since the outbreak of COVID-19, to curb the spread of the epidemic, the Chinese government has implemented unprecedented prevention interventions at the nationwide level. Currently, the outbreak in Wuhan is changing in a positive direction and has been effectively controlled. However, it is not clear what these measures were and how these measures changed to curb the outbreak of COVID-19 quickly. This study explored the characteristics and identified that China’s control strategies have changed the epidemiological curve of rapidly rising new confirmed cases of COVID-19. This study also seeks to expand the experiences and lessons from this outbreak.

**Methods:** We collected public health interventions measures from Jan 20, 2020, to 5 March 2020, and data from COVID-19 daily newly confirmed cases and daily cumulates cases to compare the control effects and changing trends. We performed a retrospective description of these intervention strategies from three stages. Besides, from the perspective of public health, the experiences and lessons exposed by this outbreak were roughly summarized.

**Results:** These non-pharmacology interventions measures adopted by the Chinese government by the instruction and spirit of President Xi Jinping were timely and efficient.

**Conclusions:** The present study was conducted to comprehensively analyze from a social epidemiology context. The results confirmed that these radical interventions taken by the Chinese government were effective, ambitious, and agile. However, we must be aware that the epidemic situation in Wuhan is still challenging.

## 1. Introduction

An unprecedented outbreak of pneumonia of unknown etiology in Wuhan, Hubei Province, China, emerged in December 2019. A novel coronavirus was identified as the causative agent and was subsequently termed SARS-CoV-2 by the World Health Organization (WHO). Epidemic disease quickly spread across 31 provinces and regions. As of 23:00 CET on March 5, a total of 98,067 new confirmed cases have been reported globally, with deaths of 3,281. A total of 17,637 cases have been confirmed in 78 countries outside China. ^[1]^ The new coronavirus outbreak has surpassed the severity of SARS in 2003.^[2, 3]^ The outbreak of virus coincided with China’s Spring Festival travel peak, and to curb the spread of the disease, the Chinese government launched unprecedented public health intervention measures, such as shutting down Wuhan transportation, extending the legal holiday, mass isolation, strict enforcement of quarantine of all contacts, and canceling all public gatherings and health public to abide by hygienic practices. To reduce the flow of people, the health monitoring of returning residents has been conducted at the nationwide level. As of 16:00 clock on February 28, Dr. Wan’nian Liang, chief of the National Health Commission’s institutional reform department and head of the Chinese expert panel, said: The virus outbreak in Wuhan is curbed basic. The number of new confirmed cases per day declined from a high peak of 3,900 cases on 13 February to 323 cases on 27 February. The number of newly cured cases has exceeded the number of newly confirmed cases for eight consecutive days since February 20. The proportion of severe cases declined from 31.6% to 22.4% from February 11 to February 27. The death rate has fallen from a peak of 9.0% on January 16th to 4.4% now. ^[4]^ As the WHO leader Bruce Aylward said at a COVID-19 news conference on February 24, “China’s made comprehensive non-pharmaceutical interventions, which have effectively prevented the transmission of the virus, providing important lessons for the global response to new virus”^[5]^. This is an unprecedented effort, and it surpasses previous efforts to combat SARS.” However, there is no doubt that no other countries can follow China’s current example.^[6]^ In the global spread background, from the perspective of social epidemiology, this paper aims to summarize these detailed intervention measures taken by the Chinese government and explain how the COVID-19 outbreak in Wuhan, Hubei Province was effectively controlled by these public health interventions measures. We also improve the public health system in the future by summarizing these lessons from this outbreak.

## 2. Methods

### 2.1. Ethical approval

This project was not applicable for institutional review board approval because it used publicly accessible information and data.

### 2.2. Data extraction and measures collection

The case report data was collected from January 11, 2020, to March 5, 2020, and the detail intervention measures conducted by China and Hubei Province were described, along with the subsequent changes in trends and analysis of its effects. Data as extracted and tabulated in Excel 2010 Microsoft and make data analysis.

**Step 1:** We extracted the data from the information reporting system of National Health Committee issued by health emergence office on January 11, 2020 - March 5, 2020, select the cumulative number of confirmed cases (Mainland in China, Hubei) and the number of daily new confirmed cases (Mainland in China, Hubei province, Wuhan city), taking the ratio of daily cumulative cases and daily new cases as the study indicators, to illustrate how these intervention measures were implemented.

**Step 2:** Relevant important documents and announcements were collected from the official website of the Chinese government and the people’s government of Hubei Province and the people’s government of Wuhan by date.

## 3. Results

### 3.1 Response measures summary and data analysis

In this paper, we made a retrospective description and summary of these inventions strategy at three stages:

**Stage 1:** During the early stage of the outbreak, before 20 January 2020, the main strategy focused on preventing the exportation of cases from Wuhan and other priority areas of Hubei Province and preventing the importation of cases by other provinces; the overall aim was to control the source infection, blocking transmission and preventing further spread. The response mechanism was initiated with multisectoral involvement in joint prevention and control measures. The Huanan Seafood Wholesale Market in the municipality of Wuhan was closed, and efforts were made to identify the zoonotic source. Information on the epidemic was sent to WHO on 3 January, and the whole genome sequence of the SARS-CoV-2 virus was shared with WHO on 10 January. Protocols for COVID-19 diagnosis and diagnosis and treatment were formulated.

Since the outbreak of COVID-19, the Chinese government has taken the most comprehensive, thorough and rigorous prevention and control measures to prevent and control the epidemic. The Central Committee of the Communist Party of China and the State Council quickly launched the national emergency response. A Central Leadership Group for Epidemic Response and the Joint Prevention and Control Mechanism of the State Council were established. President Xi Jinping personally directed and deployed the prevention and work control and requested that the prevention and control of the COVID-19 outbreak be the top priority of government at national levels.^[7]^ Prevention and control measures have been implemented rapidly, from the early stages in Wuhan and other key areas of Hubei province to the current overall national epidemic. It has been undertaken in three main phases, with important events defining those phases. COVID-19 outbreak was included in the statutory report of Class B infectious diseases and border health quarantine infectious diseases on January 20, which marked the transition from the initial partial control approach to the comprehensive adoption of various control measures by the law.

On January 22, the state council, the Communist Party of China (CPC) central committee, The official announcement that Wuhan of Hubei Province, a metropolis of 12 million people, to shut down traffic transportation to control the flow of people. On Jan 23, the authorities in Wuhan announced the lockdown of the city, shutting down all public transportation, canceling flights and trains and closing schools and factories.

On January 25, members of the standing committee of the political bureau of the CPC central committee held special meetings to restudy and redeployed and launched the prevention and control of the epidemic, especially the treatment of patients.

On January 27, Prime Minister Li Keqiang headed the Central Leading Group for Epidemic Response and went to Wuhan to inspect and coordinate the prevention and control work of relevant departments and provinces (autonomous regions and municipalities) across the country. Vice-Premier Sun Chunlan, who has been working on the front lines in Wuhan, has led and coordinated the front-line prevention and control of the outbreak.

On January 29, all provinces across China had launched the highest level of response for major public health emergencies.

On Feb 2, construction began on two temporary hospitals.

On Feb 3, medical rescue teams from 20 provinces were urgently selected to gradually transform 13 exhibition centers in Wuhan into “quadrangle hospitals” to ensure that mild patients were treated. Two days later, the first three quadrangle hospitals were opened, and the first mild cases were admitted (there were 4,250 beds in the three hospitals).On Feb 7, a temporary hospital carried out an acceptance inspection and was completed (more than 10 days) in Wuhan. On Feb 10, President Xi Jinping conducted research and guidance on the prevention and control of the epidemic in Beijing and connected by video conference with the medical staff of Wuhan Jinyintan Hospital and Wuhan Union Hospital. On Feb 12, nearly 20,000 medical staff members were sent to Wuhan, Hubei Province, and other areas for medical treatment. We organized medical staff from 19 provinces to provide support to other areas of Hubei Province.

**Stage 2:** On February 16, 2020, epidemic prevention and control actions in Wuhan and Hubei came to the most critical stages. The main strategy was to reduce the intensity of the outbreak and slow the increase in new cases. In Wuhan and other key areas of Hubei Province, the focus is on treating patients aggressively, reducing deaths and preventing export cases from exports. In other provinces, the focus is on the prevention of importation, containment of disease transmission and implementation of joint prevention and control measures. At the most critical stage of epidemic prevention and control, Wuhan was still the main battle area. As of Feb 24, there were 9 new hospitals with more than 6,960 beds, and 5,606 patients were sent to the hospital. Besides, excellent medical workers were sent to Hubei and Wuhan from all places. (1) In terms of reducing the infection rate, Hubei Province, especially Wuhan, has formulated and implemented local prevention and control measures according to the actual situation, strictly implemented the “four early stages” measures, and effectively completed the classified centralized management of “four types of people”. (2) The prevention and control measures were conducted in the community, with precise management and it was imperative to make full use of the power at the community level to curb spread from communities. (3) In terms of improving the admission rate, distinguish different situations, classify and treat all the existing patients with confirmed severe diseases, including those with clinical diagnosis, in a designated hospital, and those with confirmed mild diseases can be treated and observed at isolation places.

On Feb 16 p.m, the Hubei Province people’s government issued the three most strictest announcements within 1.5 hours. The detail orders were summarized as follows:

1. Urban and rural communities and village groups adopted closure management measures: residential areas were under the strictest closed management for 24 hours; dragnet dynamic rolling screening for all residents (only 3 days, Feb 16-19): based on the following principles: conclude the exactly cases number, do not miss a family, do not leave a person, constantly one day, to ensure full coverage; and strengthening the epidemiological investigation of the “four categories of people” (exposed infection persons, close contact of the diagnosed person, suspected cases and confirmed cases).
2. Strengthening the management of key areas. All non-essential public places shall be closed, and all mass gathering activities should be stopped.
3. Strengthening comprehensive health screening of residents: (a) the community (village), residential areas, buildings, workplaces, and other grassroots units with confirmed (including clinical diagnosis) cases of new coronavirus pneumonia shall be firmly isolated in a closed and hard manner for 14 days. (b) carry out dynamic rolling screening for all residents to ensure that “no one household was missed, no one was left behind”. Ensure all communities must have full coverage.
4. Strengthening the closure management of rural village groups requires that the natural village groups (villages) be used as the unit to implement hard isolation. Each household may send one person every three days to purchase necessary daily necessities. These detailed measures are summarized in Table 1S as supplemental files.

**Stage 3:** The third stage of the outbreak focused on reducing clusters of cases, thoroughly controlling the epidemic, and striking a balance between epidemic prevention and control, sustainable economic and social development, the unified command, standardized guidance, and scientific evidence-based policy implementation. For Wuhan and other priority areas of Hubei Province, the focus was on patient treatment and the interruption of transmission, with an emphasis on concrete steps to fully implement relevant measures for the testing, admitting and treating of all patients. Wuhan and Hubei Province continue to strictly control traffic transitions, such as controlling entrances and exits, and other provinces in China are also trying to prevent the risk of imported cases from other countries.

**Table 1.**
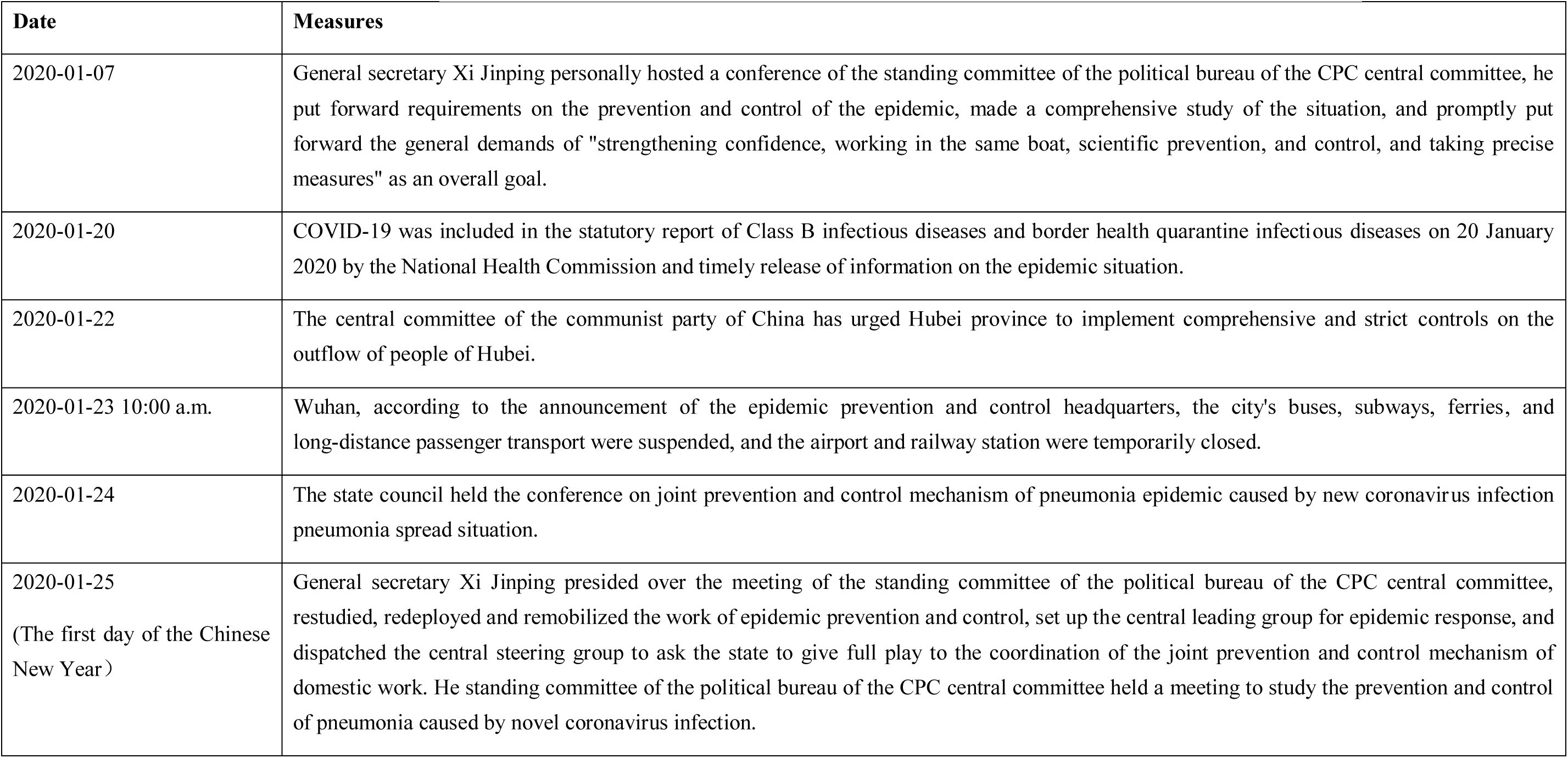
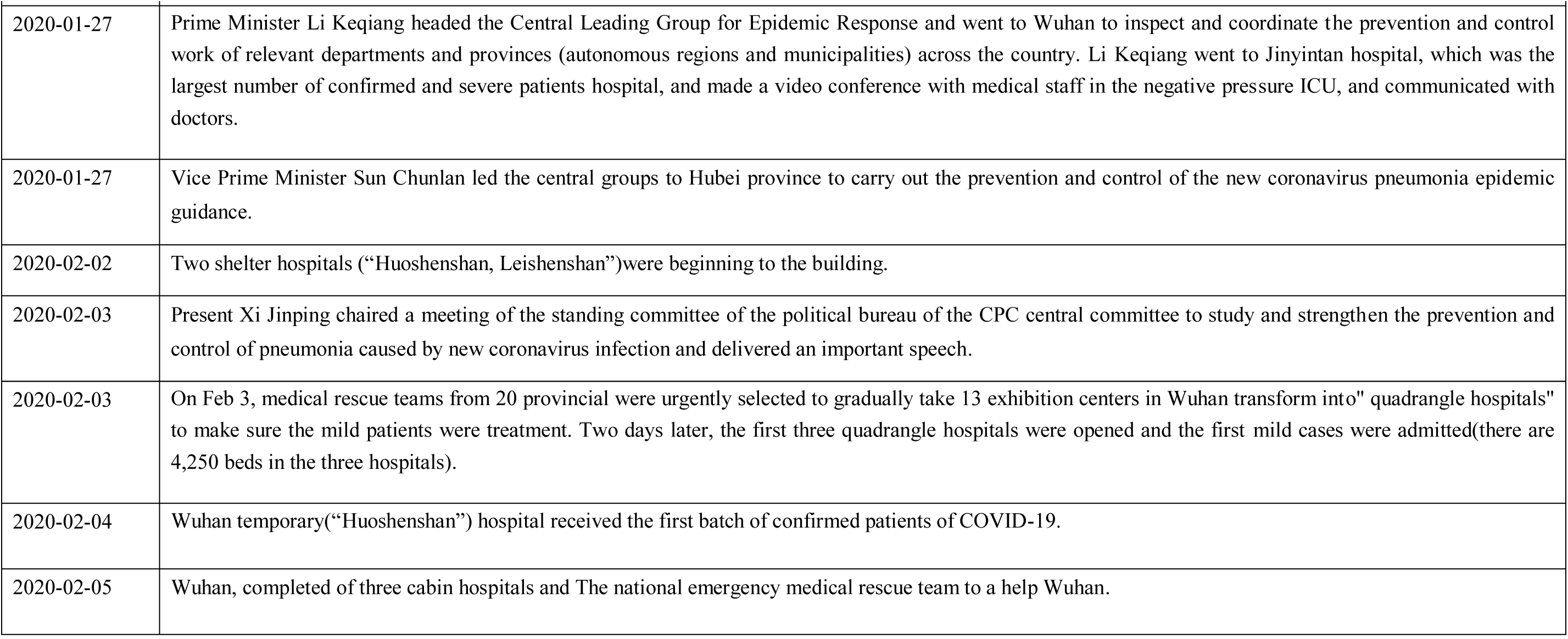
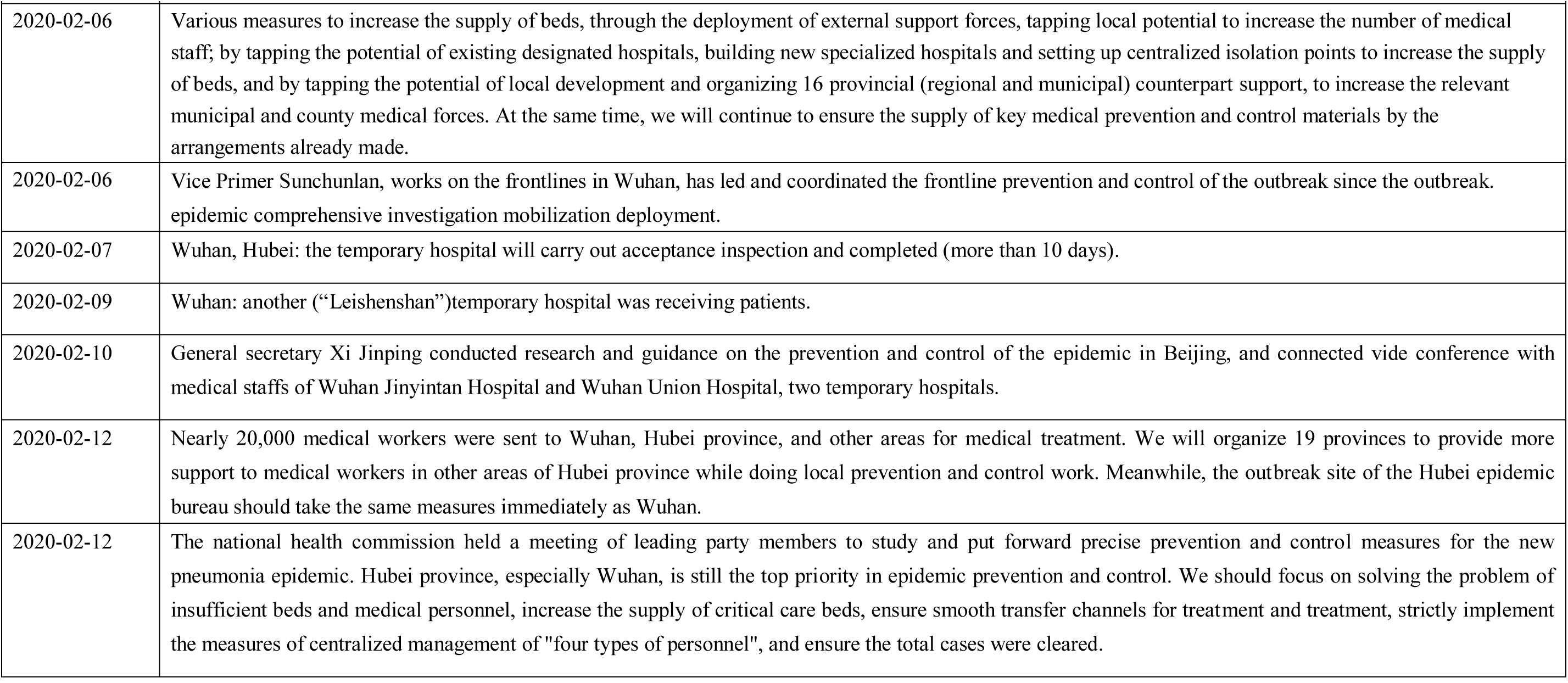
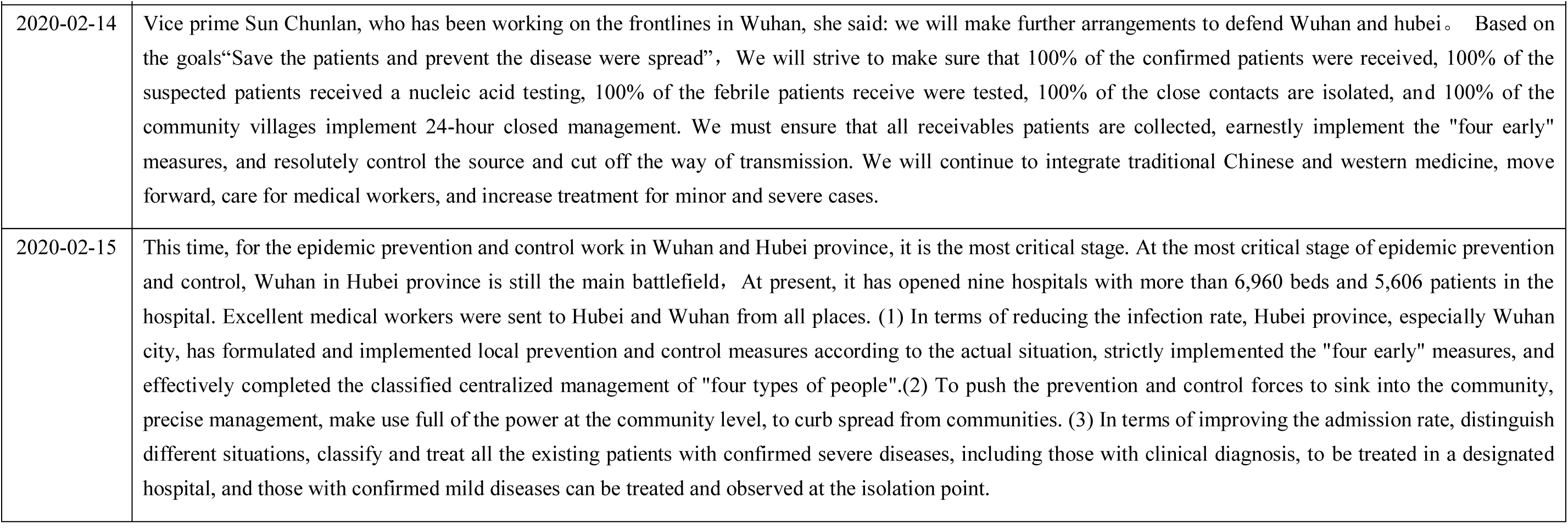
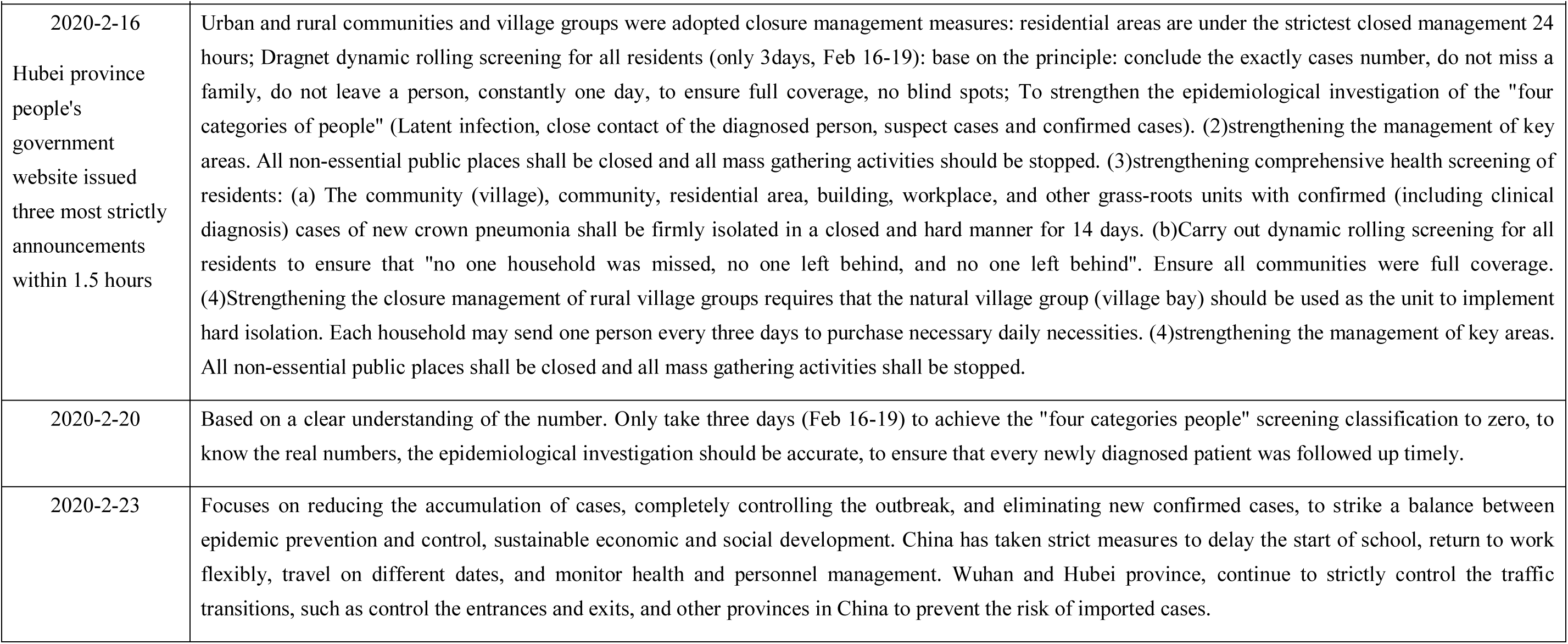
Detail public health interventions.

| Date | Measures |
| --- | --- |
| 2020-01-07 | General secretary Xi Jinping personally hosted a conference of the standing committee of the political bureau of the CPC central committee, he put forward requirements on the prevention and control of the epidemic, made a comprehensive study of the situation, and promptly put forward the general demands of "strengthening confidence, working in the same boat, scientific prevention, and control, and taking precise measures" as an overall goal. |
| 2020-01-20 | COVID-19 was included in the statutory report of Class B infectious diseases and border health quarantine infectious diseases on 20 January 2020 by the National Health Commission and timely release of information on the epidemic situation. |
| 2020-01-22 | The central committee of the communist party of China has urged Hubei province to implement comprehensive and strict controls on the outflow of people of Hubei. |
| 2020-01-23 10:00 a.m. | Wuhan, according to the announcement of the epidemic prevention and control headquarters, the city's buses, subways, ferries, and long-distance passenger transport were suspended, and the airport and railway station were temporarily closed. |
| 2020-01-24 | The state council held the conference on joint prevention and control mechanism of pneumonia epidemic caused by new coronavirus infection pneumonia spread situation. |
| 2020-01-25<br>(The first day of the Chinese New Year) | General secretary Xi Jinping presided over the meeting of the standing committee of the political bureau of the CPC central committee, restudied, redeployed and remobilized the work of epidemic prevention and control, set up the central leading group for epidemic response, and dispatched the central steering group to ask the state to give full play to the coordination of the joint prevention and control mechanism of domestic work. He standing committee of the political bureau of the CPC central committee held a meeting to study the prevention and control of pneumonia caused by novel coronavirus infection. |

| **Date** | **Measures** |
| --- | --- |
| 2020-01-27 | Prime Minister Li Keqiang headed the Central Leading Group for Epidemic Response and went to Wuhan to inspect and coordinate the prevention and control work of relevant departments and provinces (autonomous regions and municipalities) across the country. Li Keqiang went to Jinyintan hospital, which was the largest number of confirmed and severe patients hospital, and made a video conference with medical staff in the negative pressure ICU, and communicated with doctors. |
| 2020-01-27 | Vice Prime Minister Sun Chunlan led the central groups to Hubei province to carry out the prevention and control of the new coronavirus pneumonia epidemic guidance. |
| 2020-02-02 | Two shelter hospitals (“Huoshenshan, Leishenshan”)were beginning to the building. |
| 2020-02-03 | Present Xi Jinping chaired a meeting of the standing committee of the political bureau of the CPC central committee to study and strengthen the prevention and control of pneumonia caused by new coronavirus infection and delivered an important speech. |
| 2020-02-03 | On Feb 3, medical rescue teams from 20 provincial were urgently selected to gradually take 13 exhibition centers in Wuhan transform into" quadrangle hospitals" to make sure the mild patients were treatment. Two days later, the first three quadrangle hospitals were opened and the first mild cases were admitted(there are 4,250 beds in the three hospitals). |
| 2020-02-04 | Wuhan temporary(“Huoshenshan”) hospital received the first batch of confirmed patients of COVID-19. |
| 2020-02-05 | Wuhan, completed of three cabin hospitals and The national emergency medical rescue team to a help Wuhan. |

| **Date** | **Measures** |
| --- | --- |
| 2020-02-06 | Various measures to increase the supply of beds, through the deployment of external support forces, tapping local potential to increase the number of medical staff; by tapping the potential of existing designated hospitals, building new specialized hospitals and setting up centralized isolation points to increase the supply of beds, and by tapping the potential of local development and organizing 16 provincial (regional and municipal) counterpart support, to increase the relevant municipal and county medical forces. At the same time, we will continue to ensure the supply of key medical prevention and control materials by the arrangements already made. |
| 2020-02-06 | Vice Primer Sunchunlan, works on the frontlines in Wuhan, has led and coordinated the frontline prevention and control of the outbreak since the outbreak. epidemic comprehensive investigation mobilization deployment. |
| 2020-02-07 | Wuhan, Hubei: the temporary hospital will carry out acceptance inspection and completed (more than 10 days). |
| 2020-02-09 | Wuhan: another (“Leishenshan”)temporary hospital was receiving patients. |
| 2020-02-10 | General secretary Xi Jinping conducted research and guidance on the prevention and control of the epidemic in Beijing, and connected vide conference with medical staffs of Wuhan Jinyintan Hospital and Wuhan Union Hospital, two temporary hospitals. |
| 2020-02-12 | Nearly 20,000 medical workers were sent to Wuhan, Hubei province, and other areas for medical treatment. We will organize 19 provinces to provide more support to medical workers in other areas of Hubei province while doing local prevention and control work. Meanwhile, the outbreak site of the Hubei epidemic bureau should take the same measures immediately as Wuhan. |
| 2020-02-12 | The national health commission held a meeting of leading party members to study and put forward precise prevention and control measures for the new pneumonia epidemic. Hubei province, especially Wuhan, is still the top priority in epidemic prevention and control. We should focus on solving the problem of insufficient beds and medical personnel, increase the supply of critical care beds, ensure smooth transfer channels for treatment and treatment, strictly implement the measures of centralized management of "four types of personnel", and ensure the total cases were cleared. |
| 2020-02-14 | Vice prime Sun Chunlan, who has been working on the frontlines in Wuhan, she said: we will make further arrangements to defend Wuhan and hubei。 Based on the goals“Save the patients and prevent the disease were spread”, We will strive to make sure that 100% of the confirmed patients were received, 100% of the suspected patients received a nucleic acid testing, 100% of the febrile patients receive were tested, 100% of the close contacts are isolated, and 100% of the community villages implement 24-hour closed management. We must ensure that all receivables patients are collected, earnestly implement the "four early" measures, and resolutely control the source and cut off the way of transmission. We will continue to integrate traditional Chinese and western medicine, move forward, care for medical workers, and increase treatment for minor and severe cases. |
| 2020-02-15 | This time, for the epidemic prevention and control work in Wuhan and Hubei province, it is the most critical stage. At the most critical stage of epidemic prevention and control, Wuhan in Hubei province is still the main battlefield, At present, it has opened nine hospitals with more than 6,960 beds and 5,606 patients in the hospital. Excellent medical workers were sent to Hubei and Wuhan from all places. (1) In terms of reducing the infection rate, Hubei province, especially Wuhan city, has formulated and implemented local prevention and control measures according to the actual situation, strictly implemented the "four early" measures, and effectively completed the classified centralized management of "four types of people".(2) To push the prevention and control forces to sink into the community, precise management, make use full of the power at the community level, to curb spread from communities. (3) In terms of improving the admission rate, distinguish different situations, classify and treat all the existing patients with confirmed severe diseases, including those with clinical diagnosis, to be treated in a designated hospital, and those with confirmed mild diseases can be treated and observed at the isolation point. |
| 2020-2-16<br>Hubei province people's government website issued three most strictly announcements within 1.5 hours | Urban and rural communities and village groups were adopted closure management measures: residential areas are under the strictest closed management 24 hours; Dragnet dynamic rolling screening for all residents (only 3days, Feb 16-19): base on the principle: conclude the exactly cases number, do not miss a family, do not leave a person, constantly one day, to ensure full coverage, no blind spots; To strengthen the epidemiological investigation of the "four categories of people" (Latent infection, close contact of the diagnosed person, suspect cases and confirmed cases). (2)strengthening the management of key areas. All non-essential public places shall be closed and all mass gathering activities should be stopped. (3)strengthening comprehensive health screening of residents: (a) The community (village), community, residential area, building, workplace, and other grass-roots units with confirmed (including clinical diagnosis) cases of new crown pneumonia shall be firmly isolated in a closed and hard manner for 14 days. (b)Carry out dynamic rolling screening for all residents to ensure that "no one household was missed, no one left behind, and no one left behind". Ensure all communities were full coverage. (4)Strengthening the closure management of rural village groups requires that the natural village group (village bay) should be used as the unit to implement hard isolation. Each household may send one person every three days to purchase necessary daily necessities. (4)strengthening the management of key areas. All non-essential public places shall be closed and all mass gathering activities shall be stopped. |
| 2020-2-20 | Based on a clear understanding of the number. Only take three days (Feb 16-19) to achieve the "four categories people" screening classification to zero, to know the real numbers, the epidemiological investigation should be accurate, to ensure that every newly diagnosed patient was followed up timely. |
| 2020-2-23 | Focuses on reducing the accumulation of cases, completely controlling the outbreak, and eliminating new confirmed cases, to strike a balance between epidemic prevention and control, sustainable economic and social development. China has taken strict measures to delay the start of school, return to work flexibly, travel on different dates, and monitor health and personnel management. Wuhan and Hubei province, continue to strictly control the traffic transitions, such as control the entrances and exits, and other provinces in China to prevent the risk of imported cases. |

### 3.2 New confirmed cases situation

As shown in Figure 1, the highest number of confirmed cases was on 12 February. We can see that the number of new confirmed cases rose sharply before January 20, this trend almost to the third week (see Figure 1), but the epidemiological curve in Figure 1 shows that these extraordinary intervention measures directly led to flat declines or stayed at low levels from February 17 to 20 (Hubei), which indicated the situation spread of the epidemic of basic be control. The epidemiological curve showed that such measures directly lead to a flat decline or stay at a low level.

**Figure 1.**
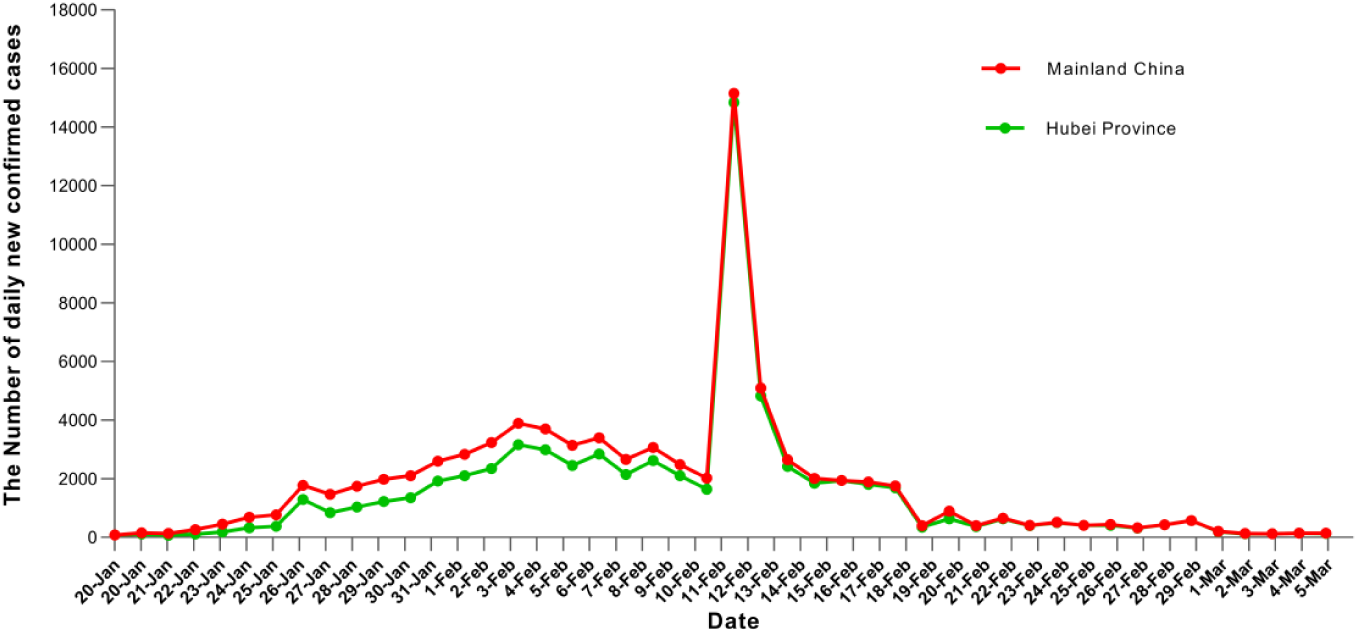
Daily Confirmed cases of COVID-19 in China and Hubei Province.

### 3.3 Change trends of new confirmed cases

Figure 2 also presents a trend that peaks near a plateau and then declines on 17 Feb 2020. Obviously, from the respectively of epidemiology, if there is such a curve, it must be due to some interventions. In this study, we believe that these positive results indicated that the new governors of Hubei Province and Wuhan, who took office from February 12, have conducted these powerful measures or actions to prevent transmission from disease rapidly.

**Figure 2.**
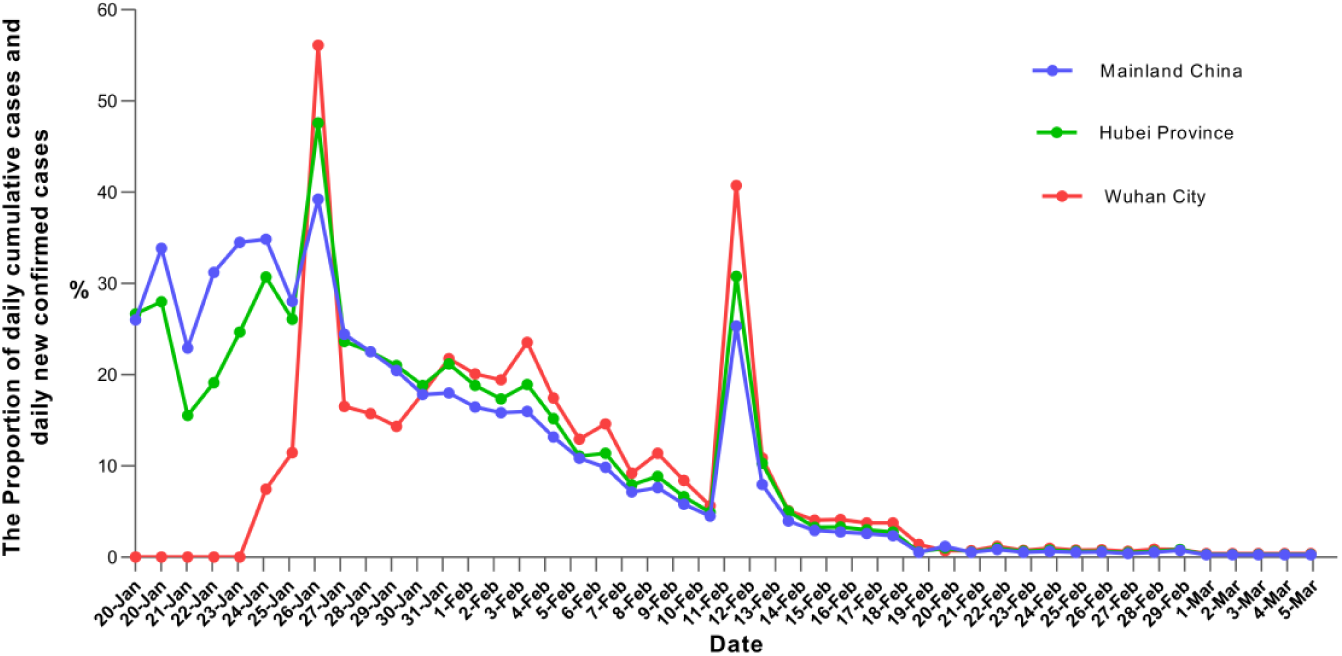
Change trends of daily newly confirmed cases and daily cumulative cases.

## 4. Discussion

In this paper, we collected these measures systematically and made respective descriptions. Our purpose is to tell the reader that COVID-19 is spreading globally at an alarming speed, and an outbreak in any environment could bring very serious consequences for each country. Some countries in Europe still take vague attitudes about this outbreak, such as “herd immunity”. People refuse to wear facemasks and so on. China’s experience has shown that non-pharmacology interventions can reduce or even stop transmission in some cases successfully. Even in the face of widespread transmission - what China’s experiences have shown was that if we keep calm, take action immediately with global, and implement systematic identification case and contact tracing, we would change the course of the epidemic, prevent people from getting sick and prevent the most vulnerable from dying.

To the best of our knowledge, the main strategy of “Four early”: “early identification, early report, early isolation and early treatment” was put forward and concentrating patients, medical experts, and medical resources into special centers to improve the rate of admission and minimize the mortality of severe patients. In Wuhan city and other priority areas of Hubei Province, the focus was on actively treating patients, reducing deaths, and preventing exports. In other provinces, the focus was on preventing importations, curbing the spread of the disease and implementing joint prevention and control measures. In this fight, medical resources from all over China have been mobilized to support the medical treatment of patients in Wuhan. A total of 343 medical teams were organized, and 42,200 medical staff members from 29 provinces and Xinjiang production and construction corps were used to assist Wuhan and Hubei Province. Less than 10 days to construct two temporary hospitals. As of 28 Feb,16 new shelter hospitals to centralize treatment patients based on the principle that all patients could be treated. It is noticed that the outbreak in China, including the hardest-hit Hubei Province, had been well under control with high-level measures for prevention and control, with temporary hospitals playing a role by separating Non-severe patients. Especially, square cabin hospitals have created a new model for China to respond to public health emergencies and crises in the future and to rapidly expand medical resources.

The Chinese government did everything possible to reserve beds, and relevant premises were repurposed to ensure that all cases could be treated on time. Wuhan and Hubei Province were treated as a priority for medical materials needs, and many provinces were organized to help one (total of 19 provinces). Because of the serious problems in virus prevention and control work in Hubei and Wuhan, the central committee of the Communist Party of China made timely adjustments to change the leading groups of the Hubei and Wuhan party committees.

In addition, new technologies such as big data and artificial intelligence are used to enhance contact tracing and focus group management and resource allocation, to identify the source of suspected cases, to conduct epidemiological investigations in the community. The data and information system for epidemic prevention and control based on the people’s public security big data system is an important “weapon” for Wuhan to win the battle of epidemic prevention and control. Additionally, the Wuhan government proposed a large step to ensure that 100% of confirmed patients should be received to the hospital, that 100% of suspected cases were undergoing nucleic acid-based testing, 100% of fever patients were detected, 100% of close contacts persons were isolation, and community villages implemented 100% 24-hours closed management. In particular, the fact of five “hundred percent” told us during February 16-19, 2020, these powerful actions to correct the statement of former Wuhan official epidemiology investigation data (98.6%)^[9]^.

Zhong Nanshan’s study team ^[8]^et al. used the population migration data on January 23, 2020, and the latest COVID-19 epidemiological data to integrate the classical infectious disease prediction model (SEIR) to predict the epidemic trend, which shows that the response measures taken by the Chinese government on January 23 effectively cubed the spread of the epidemic. If these public intervention measures had been delayed for another five days, the epidemic trend would have tripled, and the disease spread to many places and grown exponentially in one region, which is devastating for humans.

Facing a new kind of virus, there are many unknowns and no specific drugs or effective vaccines for this disease. To reduce morbidity and mortality, the Chinese government has adopted non-drug intervention measures, namely, social isolation, medical observation, contact limitation, self-protection, the combination of these traditional and modern public health intervention measures, the course of the epidemic, and the epidemic curve. As the WHO leader Bruce Aylward said at the COVID-19 news conference of February 24 in Beijing^[4]^: “This is one of the biggest successes of China’s response to the outbreak. The past month, for example, there were more than 1,800 epidemiological investigation teams in Wuhan, with at least 5 persons in each group, tracking thousands of close contacts every day. It is through painstaking efforts that the vast majority of identified close contacts were tracked down and their health monitored. These remarkable results have not come easily because of the epidemiology of these curves. Behind it in every line is an important policy decision for China’s leaders to guide the public to conform to these great decisions, which was not easy.

## 5. Limitations

In this paper, several limitations must be acknowledged. First, we did not collect suspected cases and death cases, so we didn’t discuss the measures regarding medical treatment situations in this crisis. Second, Wuhan city has been shutting down transportation more than two months, we did not discuss these mental health services about patients and their family in this outbreak, and future research needs to try to explore these topics.

## 6. Lessons and future gaps

This outbreak of COVID-19 gave us many heavy lessons. This outbreak has also sounded the alarm, revealing that China has an obvious knowledge gap in the prevention and control system and mechanism of the public health emergency management system. We summarize the following lessons based on a very basic understanding:

First, very important questions exist: Who will report when an epidemic outbreak? Where to report? Who will make these important decisions? To our knowledge, the China CDC has been established as the most effective reporting system since the SARS outbreak in 2003, which was the best reporting system in the world at that time; unfortunately, it seems this time it did not developed fully.

We believe when people are facing an outbreak of an unknown infectious disease, it would have been far better to know the first time and to report decisions to the public in a manner at the valuable time for the public health and emergency response. According to the situation in early January 2020, the data integration of multiple levels may not be realized in Wuhan. For example, the disease report data of the CDC, the diagnosis and treatment data of the medical and administrative system, the data of various research teams, etc., have not been integrated on time.

Therefore, it is necessary to build a high-level decision support center for Chinese public health security in the future to share information, comprehensively study and judge risks faced by public health security, and these value information can be timely reported to the public^[10]^.

Second, in the early outbreak, because the diagnosis process was not clear, many suspected patients had no method for confirmation and thus gave the clinicians heavy pressure, but the CDC did not develop good quality production of detection reagent in a very short time and did not ensure good production quality. In this sense, sample collection became difficult. In this process, each communication and consultation channel between the CDC and hospital did not have good collaboration in the early stage, which badly affected the early screening and diagnosis of the infected cases.

Finally, after the outbreak, with people’s panic and anxiety, residents with fever, influenza and new coronavirus infection pneumonia patients were merged into hospital fever clinics, resulting in a large number of COVID-10 patients becoming the transmission source and resulting in cross-infection, and finally giving community spread remains a huge threat. The roles of general practitioners (GPs) and the community-level health service system may not be fully developed in this outbreak.

## 7. Conclusions

The analysis results confirmed that these non-pharmacology interventions were taken by the Chinese government were effective, ambitious, and agile. However, we must be aware that the epidemic situation in Wuhan is still challenging.

## Data Availability

In this article, the datasets accessed to by National Health Committee of the People’s republic of China Official information reporting system of China. The data used to support the findings of this study are included within the article. The policy and measures issued by China government website and Hubei Province government website and some main media to support the findings of this study are included within the supplementary information file(s). The data used to support the findings of this study are available from the corresponding author upon request.

## Acknowledgments

We sincerely take the opportunity to thank anonymous reviewers for their thoughtful and meaningful comments. This study was supported by the School Foundation of Guizhou Minzu University (Grant No. GZMU([2019]YB05)).

## Abbreviations

COVID-19: novel coronavirus pneumonia
CPC: Communist Party of China
SEIR: Susceptible, Infected, Exposed, Recovered
GP: General Practitioners

## Supplement File

